# Characterization of Autonomic Symptom Burden in Long COVID: A Global Survey of 2,314 Adults

**DOI:** 10.1101/2022.04.25.22274300

**Authors:** Nicholas W. Larsen, Lauren E. Stiles, Ruba Shaik, Logan Schneider, Srikanth Muppidi, Cheuk To Tsui, Mitchell G. Miglis

**Affiliations:** Department of Neurology and Neurological Sciences, Stanford University, Palo Alto, CA; Department of Neurology, Stony Brook University Renaissance School of Medicine, Stony Brook, NY; Dysautonomia International, East Moriches, NY; Stanford Sleep Center, Department of Psychiatry and Behavioral Sciences, Stanford University, Redwood City, CA; Department of Statistics, University of Chicago, Chicago, IL

**Keywords:** COVID, autonomic, POTS, PASC, post-COVID, post-acute COVID, chronic, dysautonomia, long COVID

## Abstract

**Background:** Autonomic dysfunction is a common complication of post-acute sequalae of SARS-CoV-2 (PASC)/long COVID, however prevalence and severity rates are unknown.

**Objective:** The primary goal of this study was to assess the frequency and severity of autonomic symptoms in PASC. We also aimed to assess symptom burden in PASC though well-validated questionnaires, evaluate which pre-existing conditions are associated with an increased risk of developing autonomic dysfunction, and determine whether the severity of acute COVID-19 illness is associated with the severity of autonomic dysfunction in this population.

**Methods:** We conducted an online survey of 2,314 adults with PASC using several validated questionnaires including the COMPASS-31 to evaluate for autonomic dysfunction. We included both participants who had tested positive for COVID-19 (test-confirmed) and participants who were diagnosed with COVID-19 based on clinical symptoms alone (test-unconfirmed). Additional analyses were performed on test-confirmed participants, comparing hospitalized to non-hospitalized participants.

**Results:** 67% of PASC patients had a COMPASS-31 score >20, suggestive of moderate to severe autonomic dysfunction. COMPASS-31 scores did not differ between test-confirmed hospitalized and non-hospitalized participants (28.95±30.98 vs 26.4±28.35, p=0.06). Both hospitalized and non-hospitalized participants reported significant functional disability across all quality-of-life domains.

**Conclusions:** Moderate to severe autonomic dysfunction was seen in all PASC groups in our study, independent of hospitalization status, suggesting that autonomic dysfunction is highly prevalent in the PASC population and not necessarily dependent on the severity of acute COVID illness.

## Introduction

While the rapid growth of coronavirus disease 19 (COVID-19) cases has generally subsided globally, many individuals, regardless of symptoms and disease severity, have been left with long-lasting symptoms, a condition referred to as post-acute sequalae of SARS-CoV-2 (PASC), or long COVID. Although PASC is not universally defined, it is most often identified as symptoms of COVID-19 lasting > 30 days [1]. PASC may include a diverse constellation of symptoms, many of which appear to be autonomic in nature [2]. The most common autonomic diagnosis associated with PASC is that of postural orthostatic tachycardia syndrome (POTS), an autonomic nervous system disorder strongly associated with a preceeding viral infection [3]. In fact, many of the symptoms of PASC have been documented in individuals with POTS prior to the COVID-19 pandemic, suggesting a shared pathophysiological mechanism and prompting our interest in PASC as a framework for better understanding post-viral dysautonomia. Less commonly reported autonomic conditions associated with PASC include small fiber neuropathy [4][5], orthostatic intolerance [6], orthostatic hypotension [7], and neurally-mediated syncope [7][6]. The prevalence of POTS and other autonomic disorders in those with PASC is unknown.

While there have been online surveys of patients with PASC [4], none have specifically addressed autonomic symptom burden in relation to other features of the illness. In this context, we designed and conducted a global online survey of individuals with PASC with the goal of assessing the frequency and severity of autonomic symptoms in this population. We also aimed to: 1) assess symptom burden in PASC though well-validated questionnaires; 2) evaluate which pre-existing conditions are associated with an increased risk of developing autonomic dysfunction in PASC; and 3) determine whether the severity of acute COVID-19 illness is associated with more severe autonomic dysfunction in PASC.

## Materials and Methods

### Survey Administration

Adults ≥ 18 years of age who had self-suspected, clinician-diagnosed or test-confirmed SARS-CoV-2 infection were recruited through long COVID support groups and social media channels between October 2020 and August 2021. Self-suspected and clinician-diagnosed COVID-19 was categorized as “test-unconfirmed” and antibody, antigen and PCR-confirmed COVID-19 was categorized as “test-confirmed” for analytic purposes. All participants completed an online, English-language survey consisting of demographic information, medical history, severity of acute COVID-19 illness, and a series of validated-questionnaires. The Composite Autonomic Symptom Score-31 (COMPASS-31) was used to assess autonomic dysfunction. We also administered the Orthostatic Hypotension Questionnaire (OHQ), Fatigue Severity Scale (FSS), Epworth Sleepiness Scale (ESS), Neuropathic Pain Scale (NPS), General Anxiety Disorders Assessment (GAD-7), and the RAND 36-Item Health Survey (RAND-36). Data were collected via the online Research Electronic Data Capture (REDCap) platform. The study was approved by the Stanford University and Stony Brook University Institutional Review Boards, and all participants gave digital informed consent before starting the survey.

The COMPASS-31 is a widely-utilized patient questionnaire that provides a quantitative assessment of the severity and distribution of autonomic symptoms [5]. This questionnaire generates a weighted score from 0 to 100, and questions fall into one of six domains: orthostatic intolerance, vasomotor, secretomotor, gastrointestinal, bladder, and pupillomotor function. A COMPASS-31 score of ≥ 20 suggests moderate-to-severe autonomic dysfunction [6]. The OHQ is used to evaluate the severity of orthostatic intolerance and the functional impact of neurogenic orthostatic hypotension [7], and, by extension, other disorders of orthostatic intolerance including POTS. It consists of two components, a 6-item symptoms assessment scale and a 4-item daily activity scale. Scores range from 0 to 100, with higher scores representing higher symptom burden. The FSS is a 9-item instrument to assess the effect that fatigue has on daily functioning [8]. The FSS scores range between 9-63 points, with a score of 36 or more suggesting abnormal levels of fatigue. The ESS is a 24-point scale that quantifies the likelihood of dozing in various situations over the preceding weeks. Scores >10 suggest excessive daytime sleepiness [9]. The NPS scale measures 10 specific qualities associated with neuropathic pain. Scores range from 0 to 100, with higher scores suggesting greater disability [10]. The GAD-7 is a valid and efficient tool to screen for generalized anxiety disorder [11]. The scores range from 0-24 with the cut-offs of 5, 10, and 15 representing mild, moderate, and severe levels of anxiety. The RAND 36-Item Health Survey is a quality of life measure that incorporates eight health concepts including physical functioning, bodily pain, role limitations due to physical health problems, role limitations due to personal or emotional problems, emotional well-being, social functioning, energy/fatigue, and general health perceptions [12]. Each health concept is scored on a 0-to-100 range, with lower scores representing greater disability.

### Exclusion Criteria

In total, 4649 participants responded to the survey (figure 1). Surveys were removed from the dataset based on the following exclusion criteria: incomplete dataset, symptom duration < 30 days, symptom onset before November 2019 and age ≥ 65 years. Participants ≥ 65 years of age were excluded due to concern of survivor bias secondary to disproportionately high mortality in this age range. Ultimately, 2314 survey responses were analyzed, including 1249 test-confirmed participants and 1065 test-unconfirmed participants. Both cohorts were included but analyzed separately.

**Figure 1.**
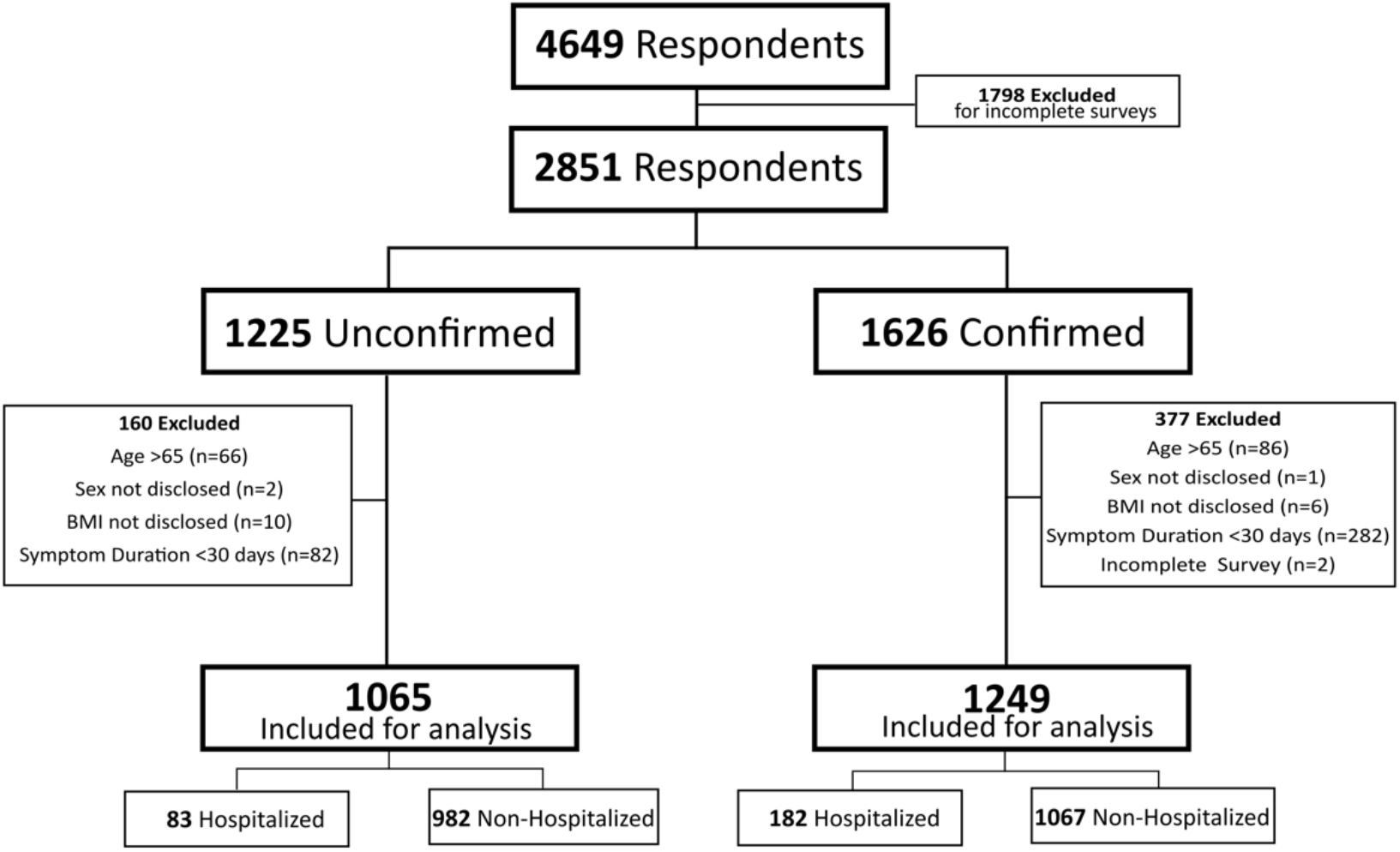
Participant flow diagram

### Statistical Analysis

A tiered approach to analyses was used to first determine comparability of the test-confirmed and test-unconfirmed groups, with subsequent analyses exploring the associations with clinical severity (i.e., hospitalized vs non-hospitalized) being assessed only in the diagnostically confirmed group. Categorical variables are presented as count and percentages, and continuous variables as mean±SD for Gaussian variables or median and interquartile range for non-Gaussian variables, as confirmed by the Shapiro-Wilk test for normality. For Gaussian variables, comparisons were performed using *t*-tests, with Cohen’s *d* for effect size estimation. For non-Gaussian variables, comparisons were performed with the Wilcoxon rank sum test, with *r* representing effect sizes. χ^2^ or Fisher’s exact test (when counts fell below 5 in any category) were used to compare categorical variables between groups, and Cramér’s *V* provided an estimation of effect size. Spearman correlation coefficients were calculated to assess strength and alignment of associations between COMPASS-31 scores and other survey measures. A correlation coefficient (r) of >0.5 or <-0.5 was interpreted as a meaningful correlation. A statistical threshold of α=0.05 was set and Bonferroni correction for multiple comparisons was performed for each major analysis. All methods were implemented in python v3.7.3[13].

## Results

### Demographics

Participants from 34 different countries responded to the survey. A majority of participants resided in the USA (85.6% test-confirmed, 68.3% test-unconfirmed), were female (86.4%, 88.4%), white (88%, 91.3%), and between the ages of 31 and 65 years (85.5%, 88.3%). A majority (61.9%) of test-confirmed participants had a body mass index (BMI) in the overweight or obese category, compared to 47.4% of test-unconfirmed participants. The test-unconfirmed group had longer duration between time of symptom onset to survey completion, with 82.4% of participants noting symptom onset ≥ 6 months prior, compared to 37.2% percent in the test-confirmed group. Most participants were never hospitalized for their acute infection (86%, 92.2%), less than 2% were admitted to the ICU and less than 1% were intubated. Complete demographics are listed in table 1.

**Table 1.** General Characteristics of Survey Respondents

| **Variable** | **Test Confirmed**<br>**(N=1249, %)** | **Test Unconfirmed**<br>**(N=1065, %)** |
| --- | --- | --- |
| **Gender** |  |  |
| Female | 1079 (86.4%) | 941 (88.4%) |
| Male | 170 (13.6%) | 124 (11.6%) |
| **Age (years)** |  |  |
| Age 18-30 | 182 (14.6%) | 126 (11.8%) |
| Age 31-45 | 494 (39.6%) | 457 (43%) |
| Age 46-65 | 573 (45.9%) | 482 (45.3%) |
| **Race/Ethnicity \*** |  |  |
| White | 1099 (88%) | 972 (91.3%) |
| Hispanic | 69 (5.5%) | 36 (3.4%) |
| Asian | 42 (3.4%) | 43 (4%) |
| Native American/Indian | 28 (2.2%) | 27 (2.5%) |
| Black | 28 (2.4%) | 14 (1.3%) |
| Middle Eastern | 11 (1 %) | 12 (1.1%) |
| Hawaiian/Pacific Islander | 6 (0.5%) | 1 (0.1%) |
| Biracial/Multiracial | 35 (2.8%) | 36 (3.4%) |
| Not disclosed/unknown | 17(1.4%) | 17 (1.6%) |
| **Average BMI\*\*** |  |  |
| Underweight (<18.5) | 41 (3.3%) | 60 (5.6%) |
| Normal (18.5-24.9) | 421 (33.7%) | 484 (45.4%) |
| Overweight (25-29.9) | 316 (25.3%) | 277 (26%) |
| Obese (30 and above) | 456 (36.6%) | 228 (21.4%) |
| **Time from symptom onset to survey completion** |  |  |
| 30 days-3 months | 426 (34.1%) | 49 (4.6%) |
| 3-6 months | 359 (28.7%) | 138 (13%) |
| 6-9 months | 352 (28.2%) | 584 (54.8%) |
| 9 months and greater | 112 (9%) | 294 (27.6%) |
| **Testing Status** |  |  |
| Self-suspected (no test) | 0 | 388 (36.4%) |
| Physician diagnosed (no test) | 0 | 677 (63.6%) |
| Symptoms only | 0 | 1065 (100%) |
| Diagnostic (RT-PCR/antigen) | 899 (72%) | 0 |
| Antibody (IgG, IgM or both) | 143 (11.4%) | 0 |
| Diagnostic and antibody | 207 (16.6%) | 0 |
| **COVID Severity\*** |  |  |
| No hospital admission | 1069 (86%) | 982 (92.2%) |
| Hospital admission | 180 (14.4%) | 83 (7.8%) |
| ICU admission | 41 (3.3 %) | 5 (0.5%) |
| Intubated | 12 (1%) | 2 (0.2%) |
| Supplemental oxygen | 134 (10.7%) | 46 (4.3%) |
| **Countries of residence** | **Respondents from 34 different countries** |  |
| USA | 1069 (85.6%) | 727 (68.3%) |
| United Kingdom | 26 (2.1%) | 128 (12%) |
| Canada | 28 (2.2%) | 103 (9.7%) |
| Germany | 27 (2.2%) | 10 (0.9%) |
| France | 5 (0.4%) | 14 (1.3%) |
| Sweden | 6 (0.5%) | 17 (1.6%) |
| Finland | 4 (0.3%) | 8 (0.7%) |
| Mexico | 5 (0.4%) | 0 (0%) |
| South Africa | 4 (0.3%) | 3 (0.3%) |
| Netherlands | 1 (0.1%) | 6 (0.6%) |
| Australia | 3 (0.2%) | 1 (0.1%) |
| Switzerland | 2 (0.2%) | 2 (0.2%) |
| Norway | 0 | 3 (0.3%) |
| India | 3 (0.2%) | 1 (0.1%) |
| Spain | 3 (0.2%) | 0 |
| No Response | 0 | 0 |
| Other | 63 (5%) | 42 (3.9%) |

\*Some respondents checked more than 1 category
\*\*Some data was incomplete (n=1234 testing confirmed, n=1049 testing unconfirmed)

**Table 2.** Medical History of Survey Respondents Prior to COVID-19

| **Variable** | **Test Confirmed**<br>**(N=1249, %)** | **Test Unconfirmed**<br>**(N=1065, %)** |
| --- | --- | --- |
| Anxiety | 412 (33%) | 252 (23.7%) |
| Depression | 299 (23.9%) | 225 (21.1%) |
| Former smoker/vaper | 255 (20.4%) | 250 (23.5%) |
| Vitamin D deficiency | 258 (20.7%) | 201 (18.9%) |
| Asthma | 256 (20.5%) | 203 (19.1%) |
| Food or environmental allergies | 195 (15.6%) | 196 (18.4%) |
| Obesity | 254 (20.3%) | 118 (11.1%) |
| High blood pressure | 212 (17%) | 109 (10.2%) |
| Autoimmune disease | 136 (10.9%) | 128 (12%) |
| Insomnia | 103 (8.2%) | 91 (8.5%) |
| Anemia | 112 (9%) | 79 (7.4%) |
| Current smoker/vaper | 68 (5.4%) | 73 (6.9%) |
| Obstructive sleep apnea | 86 (6.9%) | 54 (5.1%) |
| Neuropathy | 50 (4%) | 59 (5.5%) |
| Ehlers-Danlos syndrome | 34 (2.72%) | 67 (6.3%) |
| Diabetes | 52 (4.2%) | 29 (2.7%) |
| Cancer | 37 (3%) | 24 (2.3%) |
| Former chewing tobacco user | 10 (0.8%) | 7 (0.7%) |
| Current chewing tobacco user | 6 (0.5%) | 4 (0.4%) |
| Any form of dysautonomia | 64 (5.1%) | 88 (8.3%) |
| Postural orthostatic tachycardia syndrome | 51 (4.1%) | 73 (6.8%) |
| Vasovagal or neurocardiogenic syncope | 15 (1.2%) | 25 (2.3%) |
| Orthostatic intolerance | 9 (0.7%) | 17 (1.6%) |
| Orthostatic hypotension | 7 (0.6%) | 17 (1.6%) |
| Inappropriate sinus tachycardia | 9 (0.7%) | 13 (1.2%) |
| Autonomic neuropathy | 5 (0.4%) | 11 (1%) |
| Autonomic failure | 2 (0.2%) | 4 (0.4%) |
| Autoimmune autonomic ganglionopathy | 2 (0.2%) | 0 |
| Other types of dysautonomia | 3 (0.2%) | 6 (0.6%) |

### Medical History

The most reported pre-existing conditions were anxiety (33%, 23.7%), depression (23.9%, 21.1%), prior history of smoking or vaping (20.4%, 23.5%), vitamin D deficiency (20.7%, 18.9%), asthma (20.5%, 19.1%), food or environmental allergies (15.6%, 18.4%), obesity (20.3%, 11.1%), hypertension (17%, 10.2%) and a history of autoimmune disease (10.9%, 12%). A pre-existing diagnosis of autonomic dysfunction was reported by 5.1% of test-confirmed and 8.3% of test-unconfirmed participants, with POTS being the most commonly reported autonomic disorder (4.1%, 6.8%), followed by neurally-mediated syncope (1.2%, 2.3%), orthostatic intolerance (0.7%, 1.6%), orthostatic hypotension (0.6%, 1.6%), inappropriate sinus tachycardia (0.7%, 1.2%) and autonomic neuropathy (0.4%, 1%).

### PASC Symptom Prevalence

We grouped PASC symptoms into eight domains: respiratory, cardiovascular, gastrointestinal, pain, allergic, sensory, smell and systemic. In total, 53 different symptoms were assessed (figure 2). More than 80% of participants reported at least one symptom in each of the following domains: cardiovascular, respiratory, gastrointestinal, pain and systemic. The most reported symptoms were fatigue (89.4%, 92.4%), brain fog (81.3%, 83.7%), headache (77.9%, 80.2%), shortness of breath with exertion (73.8%, 83.8%), body aches (70.9%, 74.6%), palpitations (65%, 80.2%), lightheadedness (68.6%, 74.1%), tachycardia (62.1%, 76.7%) and difficulty sleeping (65.4%, 69.9%). Pre-syncope (48.4%, 58.3%), loss of taste (57.6%, 38.7%) and loss of smell (61.3 % 35.6%) were also commonly reported.

**Figure 2.**
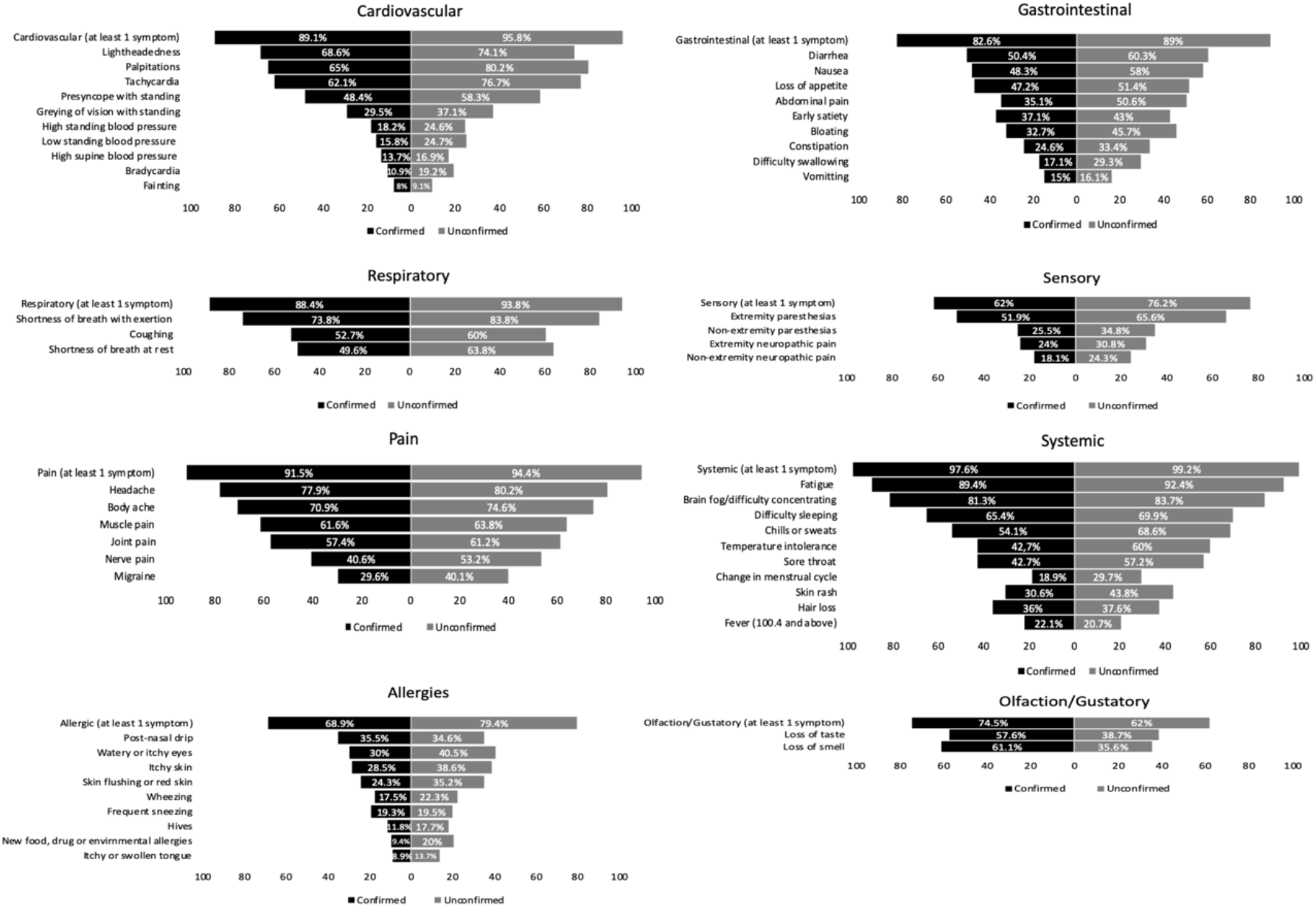
Prevalence of reported PASC symptoms

### Relationship of pre-existing conditions with autonomic symptom burden

All pre-existing conditions were associated with greater autonomic symptom burden as measured by COMPASS-31 total scores, except for high blood pressure (both groups), obesity (test-confirmed group only) and former smoker/vaper (test-unconfirmed group only) (table 3). Participants with a history of asthma (p<0.01), obesity (test-unconfirmed group only, p<0.01), vitamin D deficiency (p<0.01), autoimmune disease (p<0.01), food or environmental allergies (p<0.01), anxiety (p<0.01), depression (p<0.01), and smoking/vaping (test-confirmed group only, p<0.01) were more likely to have higher COMPASS-31 scores compared to participants who did not report these conditions. These associations were true in both test-confirmed and test-unconfirmed cohorts, unless otherwise noted.

**Table 3.** Impact of medical conditions on autonomic symptom burden

| Variable | Reported | Not Reported | T-Test Stat | Effect Size | p value | df |
| --- | --- | --- | --- | --- | --- | --- |
| High blood pressure | 212, 29.0±18.5 | 1037, 29.2±17.8 | 0.1 | -0.01 | 0.92 | 1247 |
| Asthma | 256, 33.1±18.3 | 993, 28.1±17.7 | -4.0 | 0.28 | p<0.01 | 1247 |
| Obesity | 254, 31.6±17.8 | 995, 28.5±18.0 | -2.48 | 0.17 | 0.01 | 1247 |
| Vitamin D deficiency | 258, 34.1±17.7 | 991, 27.9±17.8 | -5.0 | 0.35 | p<0.01 | 1247 |
| Autoimmune disease | 136, 35.7±18.2 | 1113, 28.4±17.8 | -4.51 | 0.41 | p<0.01 | 1247 |
| Food or environmental allergies | 195, 34.2±18.1 | 1054, 28.2±17.8 | -4.31 | 0.33 | p<0.01 | 1247 |
| Anxiety | 412, 34.2± 17.3 | 837, 26.7±17.8 | -7.10 | 0.43 | p<0.01 | 1247 |
| Depression | 299, 34.6±17.4 | 950, 27.4±17.8 | -6.06 | 0.41 | p<0.01 | 1247 |
| Smoker/Vaper | 321, 33.2±18.5 | 928, 27.8±17.6 | -4.71 | 0.3 | p<0.01 | 1247 |

Pre-existing Conditions: Test Unconfirmed Patients
| Variable | Reported | Not Reported | T-Test Stat | Effect Size | p value | df |
| --- | --- | --- | --- | --- | --- | --- |
| High blood Pressure | 109, 36.2±18.9 | 956, 33.5±17.3 | -1.51 | 0.15 | 0.13 | 1063 |
| Asthma | 203, 37.5±17.2 | 862, 32.9±17.4 | -3.38 | 0.26 | p<0.01 | 1063 |
| Obesity | 118, 38.3±18.3 | 947, 33.2±17.3 | -3.03 | 0.29 | p<0.01 | 1063 |
| Vitamin D deficiency | 201, 38.4±16.7 | 864, 32.7±17.4 | -4.24 | 0.32 | p<0.01 | 1063 |
| Autoimmune disease | 128, 38.7±17.3 | 937, 33.1±17.4 | -3.46 | 0.31 | p<0.01 | 1063 |
| Food or environmental allergies | 196, 39.1±17.4 | 869, 32.6±17.2 | -4.80 | 0.36 | p<0.01 | 1063 |
| Anxiety | 252, 39.7±17.1 | 813, 31.9±17.1 | -6.28 | 0.44 | p<0.01 | 1063 |
| Depression | 225, 39.2±17.1 | 840, 32.3±17.3 | -5.34 | 0.39 | p<0.01 | 1063 |
| Smoker/Vaper | 321, 34.7±16.9 | 744, 33.4±17.7 | -1.18 | 0.07 | 0.24 | 1063 |

### Test-Confirmed vs Test-Unconfirmed Comparison

Overall, 67 % of participants had a median COMPASS-31 score of 20 or above, suggestive of moderate to severe autonomic dysfunction. Test-unconfirmed participants had significantly higher median total COMPASS-31 scores than test-confirmed participants (test-confirmed 26.8±28.9 vs test-unconfirmed 34.7±27.8; p<0.0018) as well as significantly higher median COMPASS-31 subdomain scores in all subdomains (p<0.0018) except for secretomotor (4.29±6.43 vs 4.29±8.57; p=0.05) (table 4). Test-confirmed participants had a higher BMI than test-unconfirmed participants (27.1±10.49 vs 24.8±7.19; p<0.0018), while test-unconfirmed participants were more likely to report longer duration from symptom onset to survey completion (137±136 days vs 217±84 days; p<0.0018). Both test-confirmed and test-unconfirmed participants had elevated OHQ scores (55 [35, 73], 55[40, 70]), elevated FSS scores (54[41, 61], 56[41, 62]) and mildly elevated GAD-7 scores (8[5, 14], 7[4, 12]). The test-unconfirmed group had higher FSS (54±20 vs 56±16; p<0.0018) and NPS (12±42 vs 22±46; p<0.0018) scores than the test-confirmed group, while the test-confirmed group had elevated GAD-7 scores (8±9 vs 7±8, P<0.0018). Neither group had elevated ESS scores (8[4, 13], 8[4, 12]) suggestive of excessive daytime sleepiness. Both test-confirmed and test-unconfirmed participants had relatively low scores across all RAND subdomains, suggesting significant functional disability (table 4). The test-unconfirmed group had lower scores on the RAND-36 subdomains of physical functioning (38.89±50 vs 22±46; P<0.0018), role limitations due to physical health (0±25 vs 0±0; P<0.0018), social functioning (37.5±37.5 vs 25±37.5; P<0.0018), and general health (40±35 vs 35±25; P<0.0018).

**Table 4.**
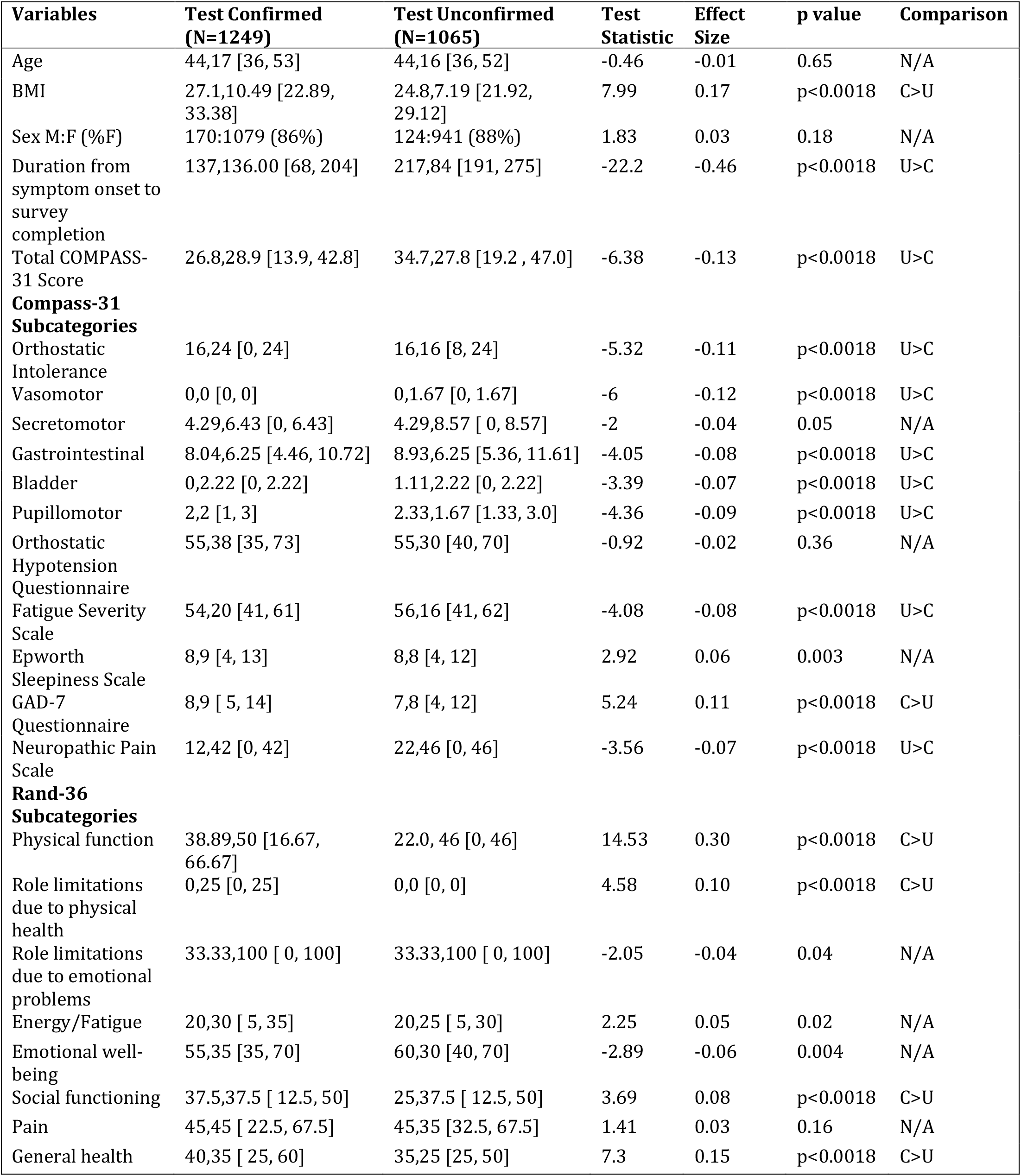
Comparison of demographic variables and symptoms scales in test-confirmed and test-unconfirmed participants.

| Variables | Test Confirmed<br>(N=1249) | Test Unconfirmed<br>(N=1065) | Test<br>Statistic | Effect<br>Size | p value | Comparison |
| --- | --- | --- | --- | --- | --- | --- |
| Age | 44,17 [36, 53] | 44,16 [36, 52] | -0.46 | -0.01 | 0.65 | N/A |
| BMI | 27.1,10.49 [22.89, 33.38] | 24.8,7.19 [21.92, 29.12] | 7.99 | 0.17 | p<0.0018 | C>U |
| Sex M:F (%F) | 170:1079 (86%) | 124:941 (88%) | 1.83 | 0.03 | 0.18 | N/A |
| Duration from symptom onset to survey completion | 137,136.00 [68, 204] | 217,84 [191, 275] | -22.2 | -0.46 | p<0.0018 | U>C |
| Total COMPASS-31 Score | 26.8,28.9 [13.9, 42.8] | 34.7,27.8 [19.2 , 47.0] | -6.38 | -0.13 | p<0.0018 | U>C |
| **Compass-31 Subcategories** |  |  |  |  |  |  |
| Orthostatic Intolerance | 16,24 [0, 24] | 16,16 [8, 24] | -5.32 | -0.11 | p<0.0018 | U>C |
| Vasomotor | 0,0 [0, 0] | 0,1.67 [0, 1.67] | -6 | -0.12 | p<0.0018 | U>C |
| Secretomotor | 4.29,6.43 [0, 6.43] | 4.29,8.57 [ 0, 8.57] | -2 | -0.04 | 0.05 | N/A |
| Gastrointestinal | 8.04,6.25 [4.46, 10.72] | 8.93,6.25 [5.36, 11.61] | -4.05 | -0.08 | p<0.0018 | U>C |
| Bladder | 0,2.22 [0, 2.22] | 1.11,2.22 [0, 2.22] | -3.39 | -0.07 | p<0.0018 | U>C |
| Pupillomotor | 2,2 [1, 3] | 2.33,1.67 [1.33, 3.0] | -4.36 | -0.09 | p<0.0018 | U>C |
| Orthostatic Hypotension Questionnaire | 55,38 [35, 73] | 55,30 [40, 70] | -0.92 | -0.02 | 0.36 | N/A |
| Fatigue Severity Scale | 54,20 [41, 61] | 56,16 [41, 62] | -4.08 | -0.08 | p<0.0018 | U>C |
| Epworth Sleepiness Scale | 8,9 [4, 13] | 8,8 [4, 12] | 2.92 | 0.06 | 0.003 | N/A |
| GAD-7 Questionnaire | 8,9 [ 5, 14] | 7,8 [4, 12] | 5.24 | 0.11 | p<0.0018 | C>U |
| Neuropathic Pain Scale | 12,42 [0, 42] | 22,46 [0, 46] | -3.56 | -0.07 | p<0.0018 | U>C |
| **Rand-36 Subcategories** |  |  |  |  |  |  |
| Physical function | 38.89,50 [16.67, 66.67] | 22.0, 46 [0, 46] | 14.53 | 0.30 | p<0.0018 | C>U |
| Role limitations due to physical health | 0,25 [0, 25] | 0,0 [0, 0] | 4.58 | 0.10 | p<0.0018 | C>U |
| Role limitations due to emotional problems | 33.33,100 [ 0, 100] | 33.33,100 [ 0, 100] | -2.05 | -0.04 | 0.04 | N/A |
| Energy/Fatigue | 20,30 [ 5, 35] | 20,25 [ 5, 30] | 2.25 | 0.05 | 0.02 | N/A |
| Emotional well-being | 55,35 [35, 70] | 60,30 [40, 70] | -2.89 | -0.06 | 0.004 | N/A |
| Social functioning | 37.5,37.5 [ 12.5, 50] | 25,37.5 [ 12.5, 50] | 3.69 | 0.08 | p<0.0018 | C>U |
| Pain | 45,45 [ 22.5, 67.5] | 45,35 [32.5, 67.5] | 1.41 | 0.03 | 0.16 | N/A |
| General health | 40,35 [ 25, 60] | 35,25 [25, 50] | 7.3 | 0.15 | p<0.0018 | C>U |

### Test Confirmed Hospitalized vs Non-Hospitalized Participants

Test-confirmed hospitalized and non-hospitalized participants both had median COMPASS-31 scores greater than 20 (hospitalized 28.95 [15.62, 46.60], non-hospitalized 26.4 [13.75, 42.10]) suggesting moderate-to-severe autonomic dysfunction, however there was no statistical difference in the severity of COMPASS-31 total (p=0.06) or subdomain scores (table 5). Test-confirmed hospitalized were older than test-confirmed non-hospitalized participants (49±15.75 vs 43±17; P<0.0018) and had a higher BMI (31.15±13.7 vs 26.62±9.66; P<0.0018). Both hospitalized and non-hospitalized participants had elevated OHQ scores (60.5 [43,77], 54 [33,71.5]), elevated FSS scores (57 [47,63], 53[39,61]) and mildly elevated GAD-7 scores (8 [5,14]], 8 [5,14]). The NPS did not differ between these two groups (p=0.007). Neither group had ESS scores suggestive of excessive daytime sleepiness (9 [5,14], 8 [4, 13]). Both hospitalized and non-hospitalized participants had relatively low scores across all RAND-36 subdomains, suggesting significant functional disability (table 5). The hospitalized group had lower subdomain scores on physical function (27.78±33.33 vs 44.44±44.45; p<0.0018), role limitations to physical health (0±0 vs 0±25; p<0.0018), social functioning (25±46.88 vs 37.5±50; p<0.0018) and general health (35±30 vs 40±30, p<0.00833). There was no difference in role limitations due to emotional problems (0±66.67 vs 33.33±100, p=0.02), energy/fatigue (20±28.75 vs 20±30, p=0.49), emotional well-being (55±33.75 vs 55±32.5, p=0.67) or pain (45±42.5 vs 45±35, p=0.005) between groups.

**Table 5.** Comparison of demographic variables and symptoms scales in test-confirmed hospitalized and test-confirmed non-hospitalized participants.

| Variables | Test Confirmed Hospitalized (N=182) | Test Confirmed Non-hospitalized (N=1067) | Test Statistic | Effect Size | p value | Comparison |
| --- | --- | --- | --- | --- | --- | --- |
| Age | 49,15.75 [ 39.25, 55] | 43,17 [ 35, 52] | 3.76 | 0.11 | p<0.0018 | CH>CNH |
| BMI | 31.15,13.7 [ 24.53, 38.23] | 26.62,9.66 [22.80, 32.46] | 5.25 | 0.15 | p<0.0018 | CH>CNH |
| Sex M:F (%F) | 31:151 (83%) | 151:928 (87%) | 1.79 | 0.04 | 0.18 | N/A |
| Duration from symptom onset to survey completion | 162,116 [91.75, 207.75] | 133,137 [ 66, 203] | 2.65 | 0.07 | 0.0078 | N/A |
| Total COMPASS-31 Score | 28.95,30.98 [ 15.62, 46.60] | 26.4,28.35 [ 13.75, 42.10] | 1.88 | 0.05 | 0.06 | N/A |
| **Compass-31 Subcategories** |  |  |  |  |  |  |
| Orthostatic Intolerance | 16,24 [ 0, 24] | 16,24 [ 0, 24] | 0.91 | 0.03 | 0.36 | N/A |
| Vasomotor | 0,1.67 [0, 1.67] | 0,0 [0, 0] | 2.66 | 0.08 | 0.008 | N/A |
| Secretomotor | 4.29,6.43 [2.14, 8.57] | 4.29,6.43 [ 0, 6.43] | 2.57 | 0.07 | 0.01 | N/A |
| Gastrointestinal | 8.04,5.36 [ 5.36, 10.72] | 8.04,6.25 [4.46, 10.72] | 1.5 | 0.04 | 0.13 | N/A |
| Bladder | 1.11,2.22 [0, 2.22] | 0,1.11 [0, 1.11] | 1.98 | 0.06 | 0.05 | N/A |
| Pupillomotor | 2.33,1.58 [ 1.42, 3.0] | 2,1.67 [ 1, 2.67] | 2.94 | 0.08 | 0.003 | N/A |
| Orthostatic Hypotension Questionnaire | 60.5,34 [ 43, 77] | 54,38.5 [33, 71.5] | 3.77 | 0.11 | p<0.0018 | CH>CNH |
| Fatigue Severity Scale | 57,16 [ 47, 63] | 53,22 [ 39, 61] | 3.99 | 0.11 | p<0.0018 | CH>CNH |
| Epworth Sleepiness Scale | 9,9 [5, 14] | 8,9 [ 4, 13] | 1.72 | 0.05 | 0.09 | N/A |
| GAD-7 Questionnaire | 8,9 [5, 14] | 8,9 [ 5, 14] | -0.09 | -0.002 | 0.93 | N/A |
| Neuropathic Pain Scale | 24.5,50.75 [ 0, 50.75] | 9,42 [ 0, 42] | 2.71 | 0.08 | 0.007 | N/A |
| **Rand-36 Subcategories** |  |  |  |  |  |  |
| Physical function | 27.78, 33.33 [ 11.11, 44.44] | 44.44,44.45[ 22.22, 66.67] | -6.47 | -0.18 | p<0.0018 | CNH>CH |
| Role limitations due to physical health | 0,0 [0, 0] | 0,25 [0, 25] | -3.4 | -0.10 | p<0.0018 | CNH>CH |
| Role limitations due to emotional problems | 0,66.67 [ 0, 66.67] | 33.33,100 [ 0, 100] | -2.38 | -0.07 | 0.02 | N/A |
| Energy/Fatigue | 20,28.75 [ 5, 33.75] | 20,30 [ 5, 35] | -0.69 | -0.02 | 0.49 | N/A |
| Emotional well-being | 55,33.75 [36.25, 70] | 55,32.5 [37.5, 70] | -0.42 | -0.01 | 0.67 | N/A |
| Social functioning | 25,46.88 [ 3.12, 50] | 37.5,50 [ 12.5, 62.5] | -3.56 | -0.10 | p<0.0018 | CNH>CH |
| Pain | 45,42.5 [22.5, 65] | 45,35 [ 32.5, 67.5] | -2.82 | -0.08 | 0.005 | N/A |
| General health | 35,30 [ 20, 50] | 40,30 [ 30, 60] | -4.25 | -0.12 | p<0.0018 | CNH>CH |

### Correlation Between COMPASS-31 and Other Survey Scores

The correlations between the total COMPASS-31 score and each of the other scales (OHQ, FSS, ESS, NPS, ESS, GAD-7 and RAND-36) can be seen in figure 3. Using a cut-off of 0.5 or -0.5, the only questionnaire with a meaningful correlation with total COMPASS-31 scores was the OHQ (test-confirmed r=0.55, test-unconfirmed; r=0.56).

**Figure 3.**
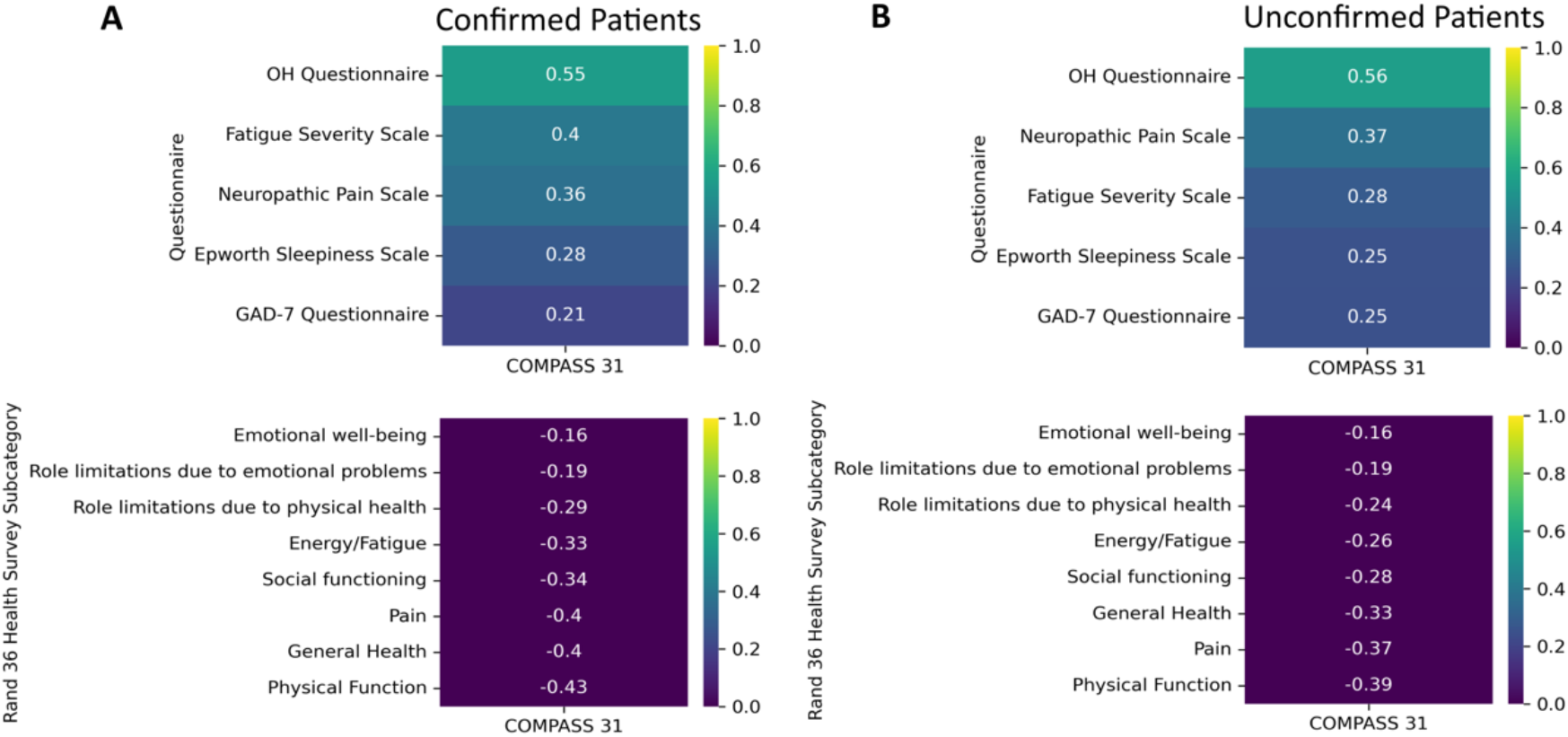
Correlations of the Fatigue Severity Scale, Neuropathic Pain Scale, Epworth Sleepiness Scale, General Anxiety Disorders Assessment, Orthostatic Hypotension Questionnaire and the Rand-36 to total COMPASS-31 scores. **\*** A lower score on the RAND 36-Item Health Survey indicates greater disability.

## Discussion

In this study we report the results of an international survey of 2,314 individuals with PASC. The percentage of female participants in this study (87%) was higher than in other studies where 68-75% percent of patients with PASC were female [14] [15]. The higher percentage of female participants in this study may also be the result of females being more likely to participate in online surveys than males [16]. Most of the PASC participants in this adult population study were between the age of 31 to 65 years (87%), which is consistent with the age distribution seen in other studies [4].

We assessed 53 different symptoms across eight symptom domains, highlighting the heterogenous nature of PASC. The most common symptoms were fatigue, brain fog, headache, shortness of breath with exertion, body aches, palpitations, lightheadedness, and tachycardia, consistent with prior studies that have noted many of the these same symptoms in PASC patients [2][17][4][18][19]. An online survey of 3,762 individuals with suspected or confirmed COVID-19 estimated the prevalence of 203 symptoms over 10 organ systems [4]. Similar to our study, some of the most commonly reported symptoms included fatigue (90-100%), brain fog (80-90%), headaches (70-80%), muscle aches (60-70%), palpitations (60-70%), dizziness or vertigo (60-70%), and tachycardia (60-70%). In our study, the test-confirmed group was more likely to report loss of taste (57.6% vs. 38.7%) and loss of smell (61.1% vs. 35.6%) than the test-unconfirmed group. We suspect that this is because individuals who experienced anosmia and ageusia were more likely to be offered testing, or more likely to pursue testing, as these symptoms were widely reported by the media and public health agencies as key indicator symptoms of acute COVID.

A pre-existing diagnosis of autonomic dysfunction was reported in 5.1% of test-confirmed and 8.3% of test-unconfirmed participants in our study, and the most common autonomic disorder reported was POTS (4.1%, 6.8%), with a prevalence that is much greater than the estimated US prevalence of 0.3-1% [20]. The most reported pre-existing conditions in our cohort were anxiety, depression, prior history of smoking or vaping, asthma, food or environmental allergies, obesity, hypertension, and a history of autoimmune disease. The prevalence of pre-COVID asthma and autoimmunity reported in our cohort are well above the rates of these medical conditions in the general US population, raising the possibility that these medical conditions could be risk factors for developing PASC [23, 24]. In comparison, the prevalence of pre-COVID anxiety and depression reported in our cohort were similar to general US population rates, suggesting that anxiety and depression are not necessarily risk factors for developing PASC [21]. All medical conditions analyzed except for hypertension (both groups), obesity (test-confirmed group), and current and former smoker/vaper (test-unconfirmed group) were associated with higher COMPASS-31 scores. The COMPASS-31 was originally designed to identify autonomic failure, which is associated with orthostatic hypotension and a reduction in sympathetic nervous system activity. Hypertension, obesity and tobacco use are all associated with sympathetic activation, [22][23][24] which may be why these conditions did not correlate with elevated COMPASS-31 scores.

Moderate to severe autonomic dysfunction was seen in all groups in our study, independent of hospitalization status, suggesting that autonomic dysfunction is highly prevalent in the PASC population and not necessarily dependent on the severity of acute COVID illness. Several other studies have evaluated autonomic dysfunction in PASC using the COMPASS-31. A cross-sectional study of 100 patients surveyed at a post-COVID unit in Colombia obtained two distinct clusters of patients based on the COMPASS-31 score [25]. Participants in the cluster with elevated COMPASS-31 scores reported more depression, chills, weakness, diarrhea, musculoskeletal pain, palpitations/tachycardia, cognitive involvement, headache, dizziness, and tinnitus. In another study from Germany, 42 patients with PASC who had persistent fatigue six months after mostly mild acute infection were assessed with the COMPASS-31 [6]. A majority (76%) had COMPASS-31 scores suggestive of moderate (score 20-40, *n* = 21) or severe (score 40-60, *n* = 11) autonomic dysfunction. Our study confirms the findings of these smaller studies and supports the concept that autonomic dysfunction is highly prevalent in PASC.

To evaluate whether the severity of acute COVID-19 infection increases the risk of developing autonomic dysfunction in PASC, we compared test-confirmed hospitalized to test-confirmed non-hospitalized participants. The hospitalized group was older and had a higher BMI than the non-hospitalized group, consistent with prior observations that increasing age and BMI are known risk factors for more severe COVID-19 infection [26]. The hospitalized group had higher scores on the OHQ (P<0.0018) and FSS (P<0.0018), as well as lower scores on several subdomains of the RAND-36, compared to the non-hospitalized group, suggesting that more severe COVID-19 illness may lead to increased orthostatic burden, worse fatigue and impaired quality of life. However, there was no difference in total COMPASS-31 scores between hospitalized and non-hospitalized participants (p=0.06), suggesting that total autonomic symptom burden in PASC is independent of the severity of the acute SARS-CoV-2 infection.

Test-unconfirmed participants had a substantially longer duration from symptom onset to survey completion compared to the test-confirmed group (P<0.0018). One explanation for this is that the test-unconfirmed group were more likely to have had COVID-19 earlier on in the pandemic, when access to testing and treatment was limited. Furthermore, hospitalized patients early in the pandemic would have been more likely to have been tested than those with milder disease, which might explain why participants in the test-unconfirmed group had lower rates of hospitalization (14.4% vs 7.8%). Interestingly, the test-unconfirmed group also had significantly greater total COMPASS-31 scores than the test-confirmed group (P<0.0018). In addition, all COMPASS-31 subdomain scores except for secretomotor were significantly greater in the test-unconfirmed group. The test-unconfirmed group also had significantly higher scores on the FSS, NPS and GAD-7, as well as lower scores multiple subdomains of the RAND-36.

There are several possible explanations for these findings. Given that the test-unconfirmed group had COVID-19 earlier in the pandemic, it is possible that participants in the test-unconfirmed group had less effective medical care. These patients may not have had access to certain therapies which are now known to improve outcomes during the acute phase of the illness. In addition, individuals who developed COVID-19 earlier in the pandemic did not have access to COVID-19 vaccines, which have been shown to reduce the acute severity of COVID-19 and reduce the risk of developing PASC ([27]). Furthermore, the acute sequelae of COVID-19 infection may have been underrecognized early in the pandemic, leading to worse outcomes. Another possibility is that test-unconfirmed participants experienced lower quality care due to a lack of a unifying diagnosis and discounting of poorly understood symptoms by clinicians, resulting in delayed access to treatment. Additionally, it is possible that different variants present different risks of developing PASC. Lastly, it is possible that some of the test-unconfirmed participants did not actually have COVID, however this would not explain why the test-unconfirmed group had higher COMPASS-31 scores. It is worth noting that 63.6 % of the test-unconfirmed participants were diagnosed with COVID-19 by a clinician.

The elevated OHQ scores seen in all groups is consistent with prior studies which have demonstrated that orthostatic intolerance syndromes are highly prevalent in PASC [28][29][30][31][32]. Similarly, the high FSS scores seen in our cohort is also consistent with prior studies that have demonstrated that fatigue is very common in PASC [18][19] [17][4]. Interestingly, none of the groups had excessively high ESS scores, suggesting that fatigue is more common than sleepiness in PASC. The mildly elevated GAD-7 scores seen in all groups is in line with data demonstrating an increase in anxiety in the general population during the pandemic [21]. All groups reported substantial impairment across all subdomains of the RAND-36, highlighting the significant impact that PASC has on quality of life, which is particularly profound in the domains of “role limitations to physical health” and “role limitations to emotional problems.”

Of all the questionnaires evaluated in this study, the only questionnaire that was strongly correlated with COMPASS-31 scores was the OHQ. It is not surprising that the OHQ scores correlated with higher COMPASS-31 scores given that disorders of orthostatic intolerance such as POTS and OH are the most common forms of dysautonomia presenting in PASC [2]. The FSS, ESS, and GAD-7 were likely not strongly associated with autonomic dysfunction because fatigue, sleepiness, and anxiety can occur, but are not specific to autonomic dysfunction.

We were surprised that the RAND-36 subdomains did not have a stronger correlation with COMPASS-31 scores. We expected that the presence of autonomic dysfunction in PASC would have had a greater impact on quality of life. These findings do not exclude the possibility that autonomic dysfunction in PASC is associated with a reduced quality of life, but rather suggest that PASC, even in the absence of autonomic dysfunction, has a very high burden of disability.

The limitations of our study include the retrospective nature, which could lead to participants either overreporting or underreporting symptoms due to recall bias. In addition, the study population was limited to those who were English speakers and had access to the PASC support groups and social media channels that our study utilized for recruitment. Finally, lack of confirmed-positive COVID test results in all patients represents a limitation inherent to the design of our survey-based study, however this is counterbalanced by the benefits of utilizing an online survey to access a large, global population of PASC patients, as well as the opportunity to include those infected early in the pandemic without access to COVID testing.

In conclusion, this is the largest study to date that has utilized validated autonomic questionnaire scores to demonstrate that autonomic dysfunction is highly prevalent in PASC. COVID severity did not correlate with the degree of autonomic dysfunction in our cohort, suggesting that even mild cases of COVID can result in significant alterations of autonomic function. Given the role of the autonomic nervous system in regulating immune function and coagulation pathways, both of which appear to play a role in long COVID [33] [34], future studies should focus on mechanisms of autonomic dysfunction in PASC, their relationship to immune and coagulation biomarkers, and potential interventions that can improve autonomic function.

## Data Availability

All data produced in the present study are available upon reasonable request to the authors.

## Acknowledgements/Contributors

We acknowledge Dysautonomia International who helped with the recruitment of participants.

## References

1. Nalbandian A, Sehgal K, Gupta A, et al (2021) Post-acute COVID-19 syndrome. Nat. Med. 27

2. Larsen NW, Stiles LE, Miglis MG (2021) Preparing for the long-haul: Autonomic complications of COVID-19. Auton. Neurosci. Basic Clin. 235

3. Shaw BH, Stiles LE, Bourne K, et al (2019) The face of postural tachycardia syndrome – insights from a large cross-sectional online community-based survey. J Intern Med 286:438–448. https://doi.org/10.1111/joim.12895

4. Davis HE, Assaf GS, McCorkell L, et al (2021) Characterizing long COVID in an international cohort: 7 months of symptoms and their impact. EClinicalMedicine 38:. https://doi.org/10.1016/j.eclinm.2021.101019

5. Sletten DM, Suarez GA, Low PA, et al (2012) COMPASS 31:a refined and abbreviated Composite Autonomic Symptom Score. Mayo Clin Proc 87:1196–201. https://doi.org/10.1016/j.mayocp.2012.10.013

6. Kedor C, Freitag H, Meyer-Arndt L, et al (2021) Chronic COVID-19 Syndrome and Chronic Fatigue Syndrome (ME/CFS) following the first pandemic wave in Germany – a first analysis of a prospective observational study. medRxiv

7. Kaufmann H, Malamut R, Norcliffe-Kaufmann L, et al (2012) The Orthostatic Hypotension Questionnaire (OHQ): Validation of a novel symptom assessment scale. Clin Auton Res 22:. https://doi.org/10.1007/s10286-011-0146-2

8. Krupp LB, Larocca NG, Muir Nash J, Steinberg AD (1989) The fatigue severity scale: Application to patients with multiple sclerosis and systemic lupus erythematosus. Arch Neurol 46:. https://doi.org/10.1001/archneur.1989.00520460115022

9. Johns MW (1991) A new method for measuring daytime sleepiness: the Epworth sleepiness scale. Sleep 14:540–5

10. Galer BS, Jensen MP (1997) Development and preliminary validation of a pain measure specific to neuropathic pain: The Neuropathic Pain Scale. Neurology 48:. https://doi.org/10.1212/WNL.48.2.332

11. Spitzer RL, Kroenke K, Williams JBW, Löwe B (2006) A brief measure for assessing generalized anxiety disorder: The GAD-7. Arch Intern Med 166:. https://doi.org/10.1001/archinte.166.10.1092

12. 36-Item Short Form Survey (SF-36) | RAND. https://www.rand.org/health-care/surveys_tools/mos/36-item-short-form.html. Accessed 20 Dec 2021

13. Guido van Rossum (1995) Python tutorial, Technical Report CS-R9526, Centrum voor Wiskunde en Informatica (CWI)

14. Post-COVID symptoms: Women differ distinctly from men -Mayo Clinic News Network. https://newsnetwork.mayoclinic.org/discussion/post-covid-symptoms-women-differ-distinctly-from-men/?_ga=2.164314809.749544649.1640119582-439971010.1640119582. Accessed 21 Dec 2021

15. Vanichkachorn G, Newcomb R, Cowl CT, et al (2021) Post–COVID-19 Syndrome (Long Haul Syndrome): Description of a Multidisciplinary Clinic at Mayo Clinic and Characteristics of the Initial Patient Cohort. Mayo Clin Proc 96:. https://doi.org/10.1016/j.mayocp.2021.04.024

16. Smith WG (2008) Does gender influence online survey participation? A record-linkage analysis of university faculty online survey response behavior. Eric Ed501717

17. Huang C, Huang L, Wang Y, et al (2021) 6-month consequences of COVID-19 in patients discharged from hospital: a cohort study. Lancet 397:. https://doi.org/10.1016/S0140-6736(20)32656-8

18. Tenforde MW, Kim SS, Lindsell CJ, et al (2020) Symptom Duration and Risk Factors for Delayed Return to Usual Health Among[1] M. W. Tenforde et al., “Symptom Duration and Risk Factors for Delayed Return to Usual Health Among Outpatients with COVID-19 in a Multistate Health Care Systems Network — United. MMWR Morb Mortal Wkly Rep 69:993–998

19. Garrigues E, Janvier P, Kherabi Y, et al (2020) Post-discharge persistent symptoms and health-related quality of life after hospitalization for COVID-19. J. Infect. 81

20. Dysautonomia International: http://www.dysautonomiainternational.org/page.php?ID=34. Accessed 21 Dec 2021

21. Vanderminden J, Esala JJ (2019) Beyond Symptoms: Race and Gender Predict Anxiety Disorder Diagnosis. Soc Ment Health 9:. https://doi.org/10.1177/2156869318811435

22. Grassi G (2015) Sympathetic overdrive in hypertension: clinical and therapeutic relevance. J Cardiol Pr 13:24

23. Esler M, Straznicky N, Eikelis N, et al (2006) Mechanisms of sympathetic activation in obesity-related hypertension. Hypertension 48

24. Narkiewicz K, Van De Borne PJH, Hausberg M, et al (1998) Cigarette smoking increases sympathetic outflow in humans. Circulation 98:. https://doi.org/10.1161/01.CIR.98.6.528

25. Anaya JM, Rojas M, Salinas ML, et al (2021) Post-COVID syndrome. A case series and comprehensive review. Autoimmun. Rev. 20

26. Kompaniyets L, Pennington AF, Goodman AB, et al (2021) Underlying Medical Conditions and Severe Illness Among 540,667 Adults Hospitalized With COVID-19, March 2020–March 2021. Prev Chronic Dis 18:1–13. https://doi.org/10.5888/PCD18.210123

27. Kuodi P, Gorelik Y, Zayyad H, et al (2022) Association between vaccination status and reported incidence of post-acute COVID-19 symptoms in Israel: a cross-sectional study of patients infected between March 2020 and November 2021. medRxiv 2022.01.05.22268800

28. Miglis MG, Prieto T, Shaik R, et al (2020) A case report of postural tachycardia syndrome after COVID-19. Clin. Auton. Res. 30

29. Kanjwal K, Jamal S, Kichloo A, Grubb B (2020) New-onset Postural Orthostatic Tachycardia Syndrome Following Coronavirus Disease 2019 Infection. J Innov Card Rhythm Manag 11:. https://doi.org/10.19102/icrm.2020.111102

30. Dani M, Dirksen A, Taraborrelli P, et al (2021) Autonomic dysfunction in ‘long COVID’: rationale, physiology and management strategies. Clin Med J R Coll Physicians London 21:. https://doi.org/10.7861/CLINMED.2020-0896

31. Johansson M, Ståhlberg M, Runold M, et al (2021) Long-Haul Post–COVID-19 Symptoms Presenting as a Variant of Postural Orthostatic Tachycardia Syndrome: The Swedish Experience. JACC Case Reports 3:. https://doi.org/10.1016/j.jaccas.2021.01.009

32. Rass V, Beer R, Schiefecker AJ, et al (2020) Neurological Outcome and Quality of Life Three Months after COVID-19: A Prospective Observational Cohort Study. SSRN Electron J. https://doi.org/10.2139/ssrn.3733679

33. Utrero-Rico A, Ruiz-Ruigómez M, Laguna-Goya R, et al (2021) A short corticosteroid course reduces symptoms and immunological alterations underlying long-COVID. Biomedicines 9:. https://doi.org/10.3390/biomedicines9111540

34. Pretorius E, Vlok M, Venter C, et al (2021) Persistent clotting protein pathology in Long COVID/Post-Acute Sequelae of COVID-19 (PASC) is accompanied by increased levels of antiplasmin. Cardiovasc Diabetol 20:. https://doi.org/10.1186/s12933-021-01359-7

